# Detection of SARS-CoV-2 variants by Abbott molecular, antigen, and serological tests

**DOI:** 10.1101/2021.04.24.21256045

**Authors:** Mary A Rodgers, Rahul Batra, Luke B Snell, David Daghfal, Richard Roth, Shihai Huang, Stephen Kovacs, Gaia Nebbia, Sam Douthwaite, Gavin A Cloherty

**Author notes:** Corresponding author: Mary A Rodgers, 100 Abbott Park Rd, Abbott Park, IL 60064.

## Abstract

**Background:** Viral diversity presents an ongoing challenge for diagnostic tests, which need to accurately detect all circulating variants. The Abbott Global Surveillance program monitors severe acute respiratory syndrome coronavirus-2 (SARS-CoV-2) variants and their impact on diagnostic test performance.

**Objectives:** To evaluate the capacity of Abbott molecular, antigen, and serologic assays to detect the SARS-CoV-2 B.1.1.7, B.1.351 and the P.1 variants.

**Study design:** Virus variant culture stock dilutions (B.1.1.7, BEI NR-54011; B.1.351, BEI NR-54008 and 54009; P.1, BEI NR-54982) and clinical samples from patients with confirmed B.1.1.7 variant infection were run on the Abbott ID NOW COVID-19, *m*2000 RealTi*m*e SARS-CoV-2, Alinity m SARS-CoV-2, and Alinity m Resp-4-Plex molecular assays; the BinaxNOW COVID-19 Ag Card and Panbio COVID-19 Ag Rapid Test Device; and the ARCHITECT/Alinity i SARS-CoV-2 IgG and AdviseDx IgM assays, Panbio COVID-19 IgG assay, and ARCHITECT/Alinity i AdviseDx SARS-CoV-2 IgG II assay.

**Results:** Cultured virus stocks and B.1.1.7 clinical samples were detected with molecular, antigen, and serologic assays in the expected ranges, confirming *in silico* predictions. The ratio between genome equivalents (GE) and calculated median tissue culture infectious dose (TCID_50_) were 31-to 83-fold higher for B.1.1.7 cultures compared to B.1.351 and P.1 cultures, demonstrating that GE are more consistent units between cultures than TCID_50_.

**Conclusions:** Abbott molecular and antigen assays effectively detect B.1.1.7, B.1.351, and P.1 variant infections and Abbott serologic assays detect B.1.1.7 antibodies in patient sera. Future studies with SARS-CoV-2 virus cultures should use quantitative viral load values to compare detection of variants.

**Highlights:**

- Abbott SARS-CoV-2 molecular and antigen assays detect B.1.1.7, B.1.351, and P.1 variants
- Abbott SARS-CoV-2 antibody assays detect B.1.1.7 antibodies in recovered patient sera
- Quantitation of viral load in genome equivalents allows comparison of assay performance

## 1. Background

As viruses continue to evolve, diagnostic tests must keep pace to ensure accurate detection of all circulating variants. While assay design can mitigate the impact of viral diversity, assay performance must be continually monitored through variant testing and molecular surveillance. To meet this challenge, the Abbott Global Surveillance program has been tracking the sequences of severe acute respiratory syndrome coronavirus-2 (SARS-CoV-2) variants that have emerged throughout the pandemic, including three major lineages that have spread globally: B.1.1.7, B.1.351, and P.1. The B.1.1.7 lineage was first identified in the United Kingdom and carries spike mutations that have been linked to increased transmissibility, including N501Y [1-3]. The B.1.351 lineage was first identified in South Africa and has since spread to more than a dozen countries, with initial reports indicating that this variant can escape neutralizing antibodies [4-6]. The P.1 variant was first reported in Japan as a branch of the B.1.128 lineage, but was subsequently traced back to Brazil. The P.1 variant contains similar mutations in the spike gene as the B.1.1.7 and B.1.351 variants, suggesting a convergence of SARS-CoV-2 spike mutations that may increase transmissibility and risk of re-infection [7]. While spike gene mutations primarily define all three of the major variant lineages, the presence of additional mutations throughout the genome warrant further examination for potential impact on diagnostic assay performance.

## 2. Objective

The goal of this study was to evaluate the performance of Abbott SARS-CoV-2 molecular, antigen, and serological assays to detect SARS-CoV-2 B.1.17, B.1.351, and P.1 variants and B.1.1.7 antibodies.

## 3. Study design

B.1.1.7, B.1.351, and P.1 lineage sequences were obtained from GISAID [8] for *in silico* comparison to assay target sequences. Virus cultures for the B.1.1.7 variant (BEI NR-54011) [9], the B.1.351 variant (BEI NR-54008, NR-54009) [10, 11], and the P.1 variant (BEI NR-54982) [12] were heat inactivated at 65°C for 30 minutes and diluted for testing with the Abbott ID NOW COVID-19, *m*2000 RealTi*m*e SARS-CoV-2, Alinity m SARS-CoV-2, and Alinity m Resp-4-Plex molecular assays; and the BinaxNOW COVID-19 Ag Card and Panbio COVID-19 Ag Rapid Test Device. Antigen testing was performed by pipetting the virus stock dilution directly onto a sterile swab and then following the manufacturer’s instructions for use.

Anonymized leftover patient nasopharyngeal swab samples in viral transport media (VTM), collected at the point of discard from inpatients at Guys’ and St Thomas’ Hospital in London (UK Research Ethics Committee 20/SC/0310), were sequenced using the ARTIC protocol [13] on Nanopore instruments to determine infection with the B.1.1.7 variant, then run neat or diluted 5-62.5x before molecular testing on the Abbott ID NOW COVID-19, *m*2000 RealTi*m*e SARS-CoV-2, Alinity m SARS-CoV-2, and Alinity m Resp-4-Plex. Matched serum samples from the same patients were used for serologic testing with the ARCHITECT/Alinity i SARS-CoV-2 IgG and AdviseDx IgM assays, Panbio COVID-19 IgG assay, and ARCHITECT/Alinity i AdviseDx SARS-CoV-2 IgG II assay. All molecular and serological assays were performed per the manufacturer’s instructions for use.

## 4. Results

Initial *in silico* examination of B.1.1.7, B.1.351, or P.1 sequences revealed no lineage-defining mutations of concern for the performance of Abbott assays targeting the RDRP region (ID NOW COVID-19, *m*2000 RealTi*m*e SARS-CoV-2, Alinity m SARS-CoV-2, Alinity m Resp-4-Plex), nucleocapsid region (*m*2000 RealTi*m*e SARS-CoV-2, Alinity m SARS-CoV-2, Alinity m Resp-4-Plex, BinaxNOW COVID-19 Ag Card, Panbio COVID-19 Ag Rapid Test Device, ARCHITECT/Alinity i SARS-CoV-2 IgG, and Panbio COVID-19 IgG), and spike region (ARCHITECT/Alinity i AdviseDx SARS-CoV-2 IgM, ARCHITECT/Alinity i AdviseDx SARS-CoV-2 IgG II). To evaluate these predictions for Abbott’s molecular and antigen assays, dilution series of heat-inactivated B.1.1.7 (BEI NR-54011) [9], B.1.351 (BEI NR-54008, NR-54009) [10, 11], and P.1 (BEI NR-54982) [12] virus cultures were tested on each assay. Multiple dilutions were detected with *m*2000, Alinity m, ID NOW, BinaxNOW, and Panbio Ag assays in the expected ranges previously observed with other strains (Tables 1-3) [14, 15]. These results confirm the *in silico* predictions and are consistent with recent Panbio Ag evaluations of the B.1.1.7 and B.1.351 lineages [16, 17].

**Table 1.**
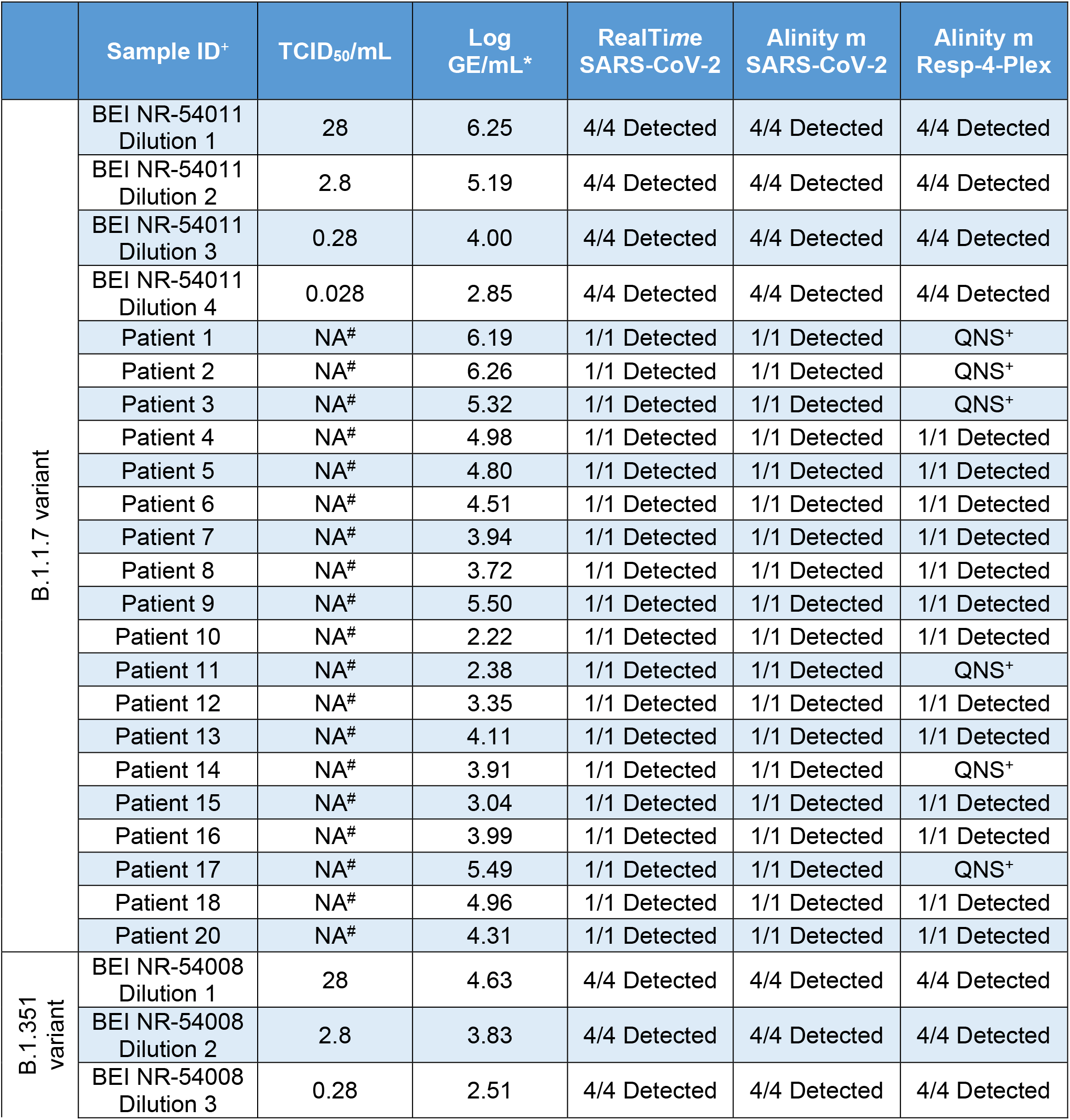
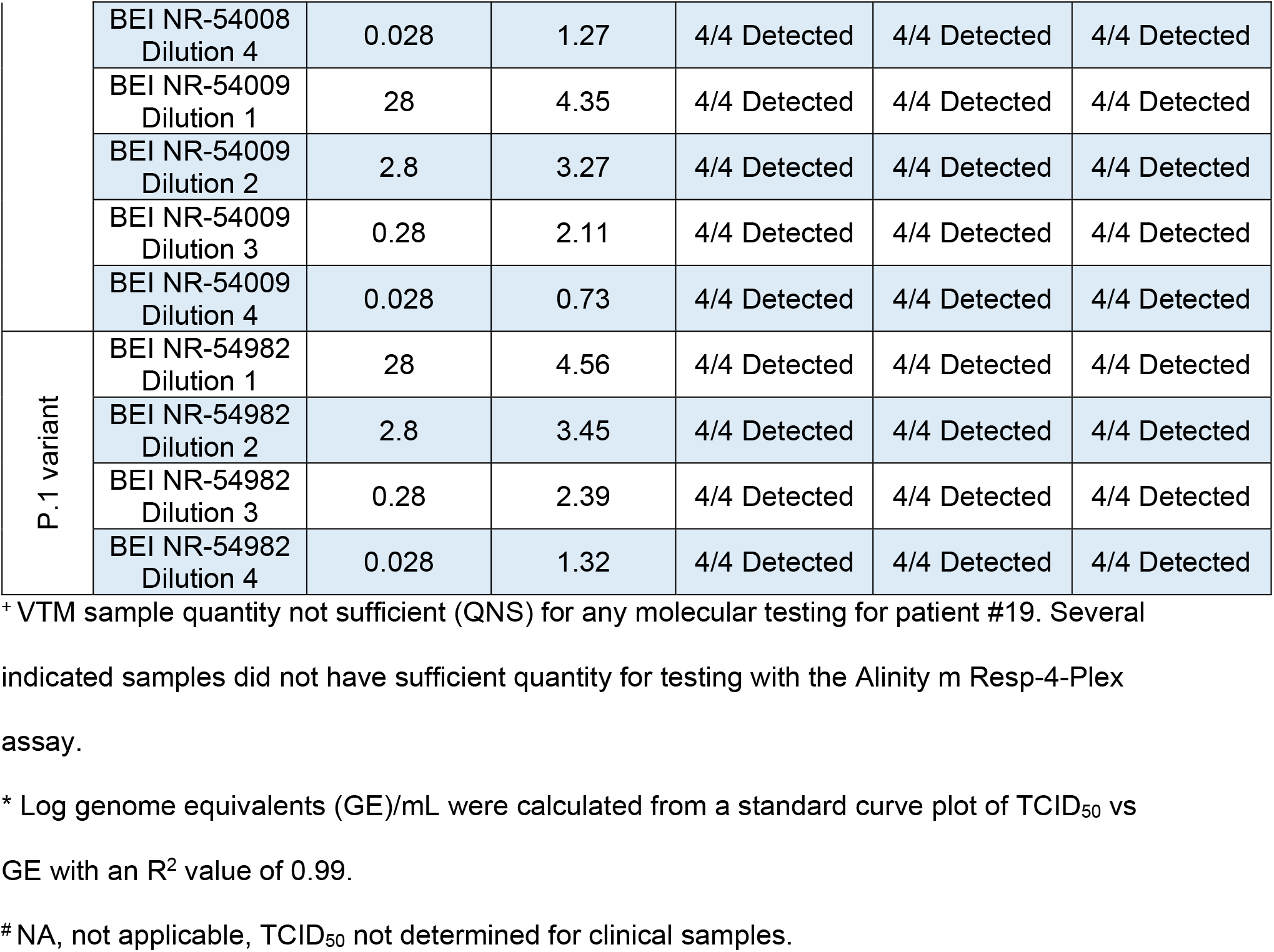
*m*2000 and Alinity m SARS-CoV-2 Assay Results With B.1.1.7, B.1.351, and P.1 Virus Culture Dilutions and B.1.1.7 Clinical Samples

**Table 2.**
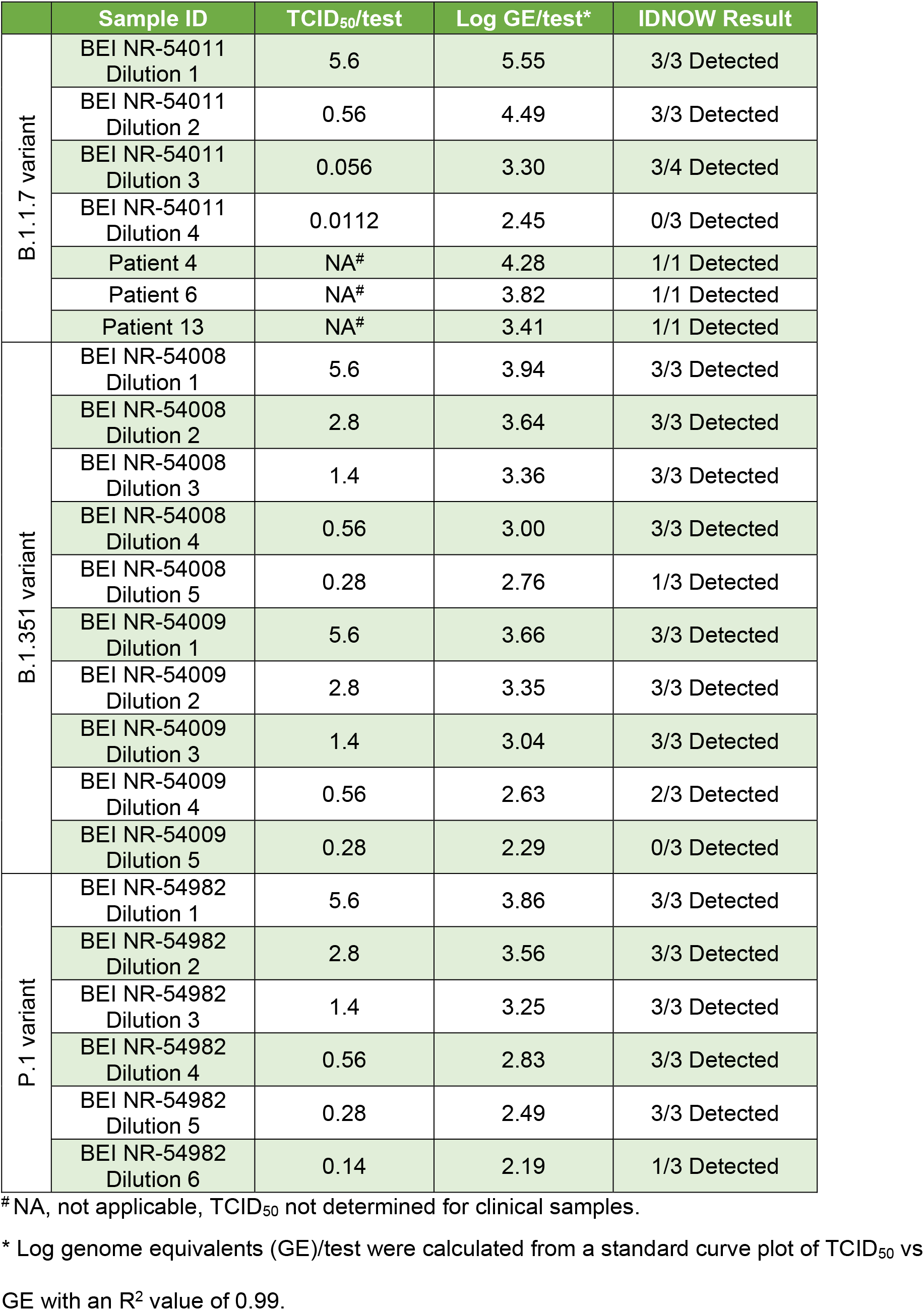
Rapid Molecular Assay Results With ID NOW COVID-19 with B.1.1.7, B.1.351, and P.1 Virus Culture Dilutions and B.1.1.7 Clinical Samples

**Table 3.** Rapid Antigen Assay Results with Virus Culture Dilutions for B.1.1.7 (BEI NR-54011), B.1.351 (BEI NR-54008 and 54009), and P.1 (NR-54982)

|  | Culture | TCID<sub>50</sub>/test | Log GE/test* | BinaxNOW Result | Panbio Ag Result |
| --- | --- | --- | --- | --- | --- |
| B.1.1.7 variant | Neat | 560 | 7.56 | 1/1 Positive | 1/1 Positive |
|  | BEI NR-54011 Dilution 1 | 56 | 6.56 | 4/4 Positive | 3/3 Positive |
|  | BEI NR-54011 Dilution 2 | 28 | 6.26 | 4/4 Positive | 3/3 Positive |
|  | BEI NR-54011 Dilution 3 | 14 | 5.96 | 3/4 Positive | 3/4 Positive |
| B.1.351 variant | BEI NR-54008 Dilution 1 | 1160 | 6.25 | 3/3 Detected | 3/3 Detected |
|  | BEI NR-54008 Dilution 2 | 928 | 6.15 | 3/3 Detected | 3/3 Detected |
|  | BEI NR-54008 Dilution 3 | 560 | 5.93 | 6/6 Detected | 3/3 Detected |
|  | BEI NR-54008 Dilution 4 | 56 | 4.93 | 0/3 Detected | 0/3 Detected |
|  | BEI NR-54009 Dilution 1 | 1160 | 5.98 | 3/3 Detected | 3/3 Detected |
|  | BEI NR-54009 Dilution 2 | 928 | 5.88 | 3/3 Detected | 3/3 Detected |
|  | BEI NR-54009 Dilution 3 | 560 | 5.66 | 6/6 Detected | 3/3 Detected |
|  | BEI NR-54009 Dilution 4 | 56 | 4.66 | 3/3 Detected | 0/3 Detected |
| P.1 variant | BEI NR-54982 Dilution 1 | 1160 | 6.18 | 3/3 Detected | 3/3 Detected |
|  | BEI NR-54982 Dilution 2 | 928 | 6.08 | 3/3 Detected | 3/3 Detected |
|  | BEI NR-54982 Dilution 3 | 560 | 5.86 | 3/3 Detected | 3/3 Detected |
|  | BEI NR-54982 Dilution 4 | 280 | 5.56 | 3/3 Detected | 3/3 Detected |
|  | BEI NR-54982 Dilution 5 | 56 | 4.86 | 0/3 Detected | 0/3 Detected |

\* Log genome equivalents (GE)/test were calculated from a standard curve plot of TCID<sub>50</sub> vs GE with an R<sup>2</sup> value of 0.99.

Considerable variability was observed in the ratio between genome equivalents (GE) quantitated with a standard curve and the calculated median tissue culture infectious dose (TCID_50_) for each virus isolate stock (Table 4). The GE/TCID_50_ ratios were 23 to 102-fold higher on average for the B.1.1.7 culture than either of the B.1.351 stocks or the P.1 stock. This difference should be taken into account when evaluating assay performance with SARS-CoV-2 virus cultures and highlights the need for standardized comparisons in units that are independent of virus culture conditions, which can vary between preparations.

**Table 4.**
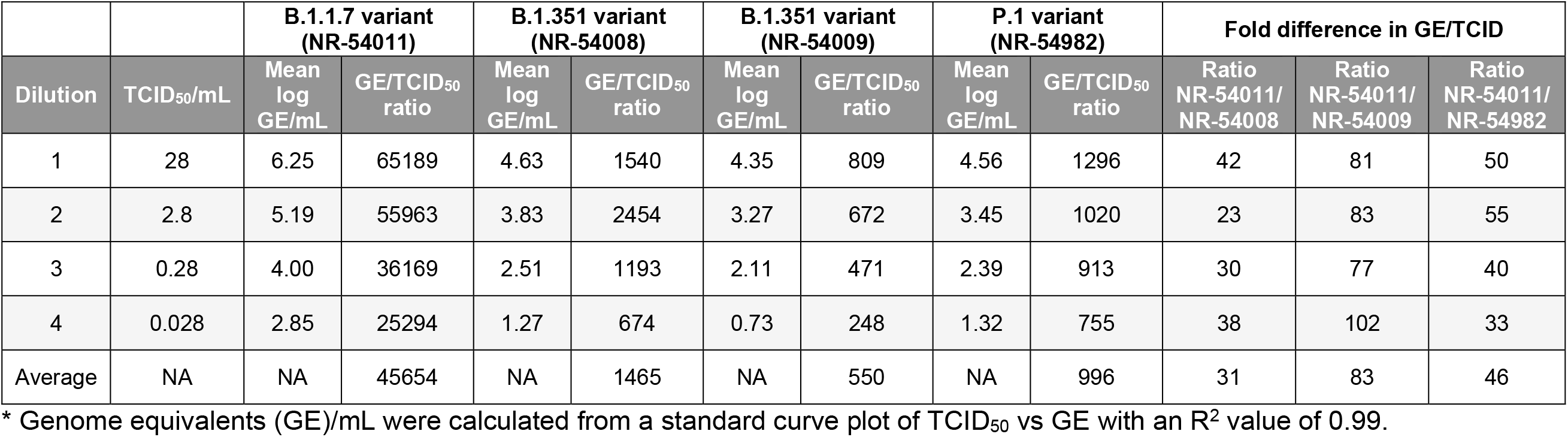
Comparison of GE*/TCID_50_ Ratios

Further evaluation was performed with leftover nasopharyngeal swab samples in VTM and matched serum samples from 20 patients with sequence confirmed B.1.1.7 infections. Nineteen VTM samples were run on the *m*2000 and both Alinity m SARS-CoV-2 assays; the sample volume from patient #19 was not sufficient for any molecular testing. The B.1.1.7 variant was detected in all 19 samples by the *m*2000 and both Alinity m assays and was detected in all 13 samples with sufficient volume by the Alinity Resp-4-Plex assay (Table 1). Three samples had sufficient volume for testing with ID NOW; the B.1.1.7 variant was detected in all three (Table 2).

Serological assays were run with serum collected from the same B.1.1.7 patients 15-26 days post symptom onset (Table 4). Notably, three immune-compromised patients (#2, #4, and #10) were included in the panel. Antibodies for patients #2 and #4 were detected by four assays (ARCHITECT and Alinity i IgM and IgG II), but not by ARCHITECT, Alinity i, or Panbio IgG assays. Patient #10 had cancer and did not have a detectable antibody response at 15 days post symptom onset by any assay. Excluding immune-compromised patients, antibodies were detected in 100% (17/17) of patients by ARCHITECT/Alinity i IgM and IgG II assays and were detected in 94% (16/17) of patients by ARCHITECT/Alinity i, and Panbio IgG assays (Table 5).

**Table 5.** SARS-CoV-2 Serology Assay Results with B.1.1.7 Clinical Samples

| Sample ID | ARCH IgM (Index)<sup>†</sup> | Alinity IgM (Index)<sup>†</sup> | ARCH IgG (Index)<sup>†</sup> | Alinity IgG (Index)<sup>†</sup> | ARCH IgG II (AU/mL)<sup>†</sup> | Alinity IgG II (AU/mL)<sup>†</sup> | Panbio IgG Result | Days post symptom onset |
| --- | --- | --- | --- | --- | --- | --- | --- | --- |
| Patient 1 | 18.36 | 18.68 | 6.76 | 6.97 | 13359.5 | 11595.4 | Positive | 26 |
| Patient 2 | 1.22 | 1.26 | 0.22 | 0.22 | 178.5 | 189.5 | Negative | 20 |
| Patient 3 | 11.68 | 12.05 | 5.62 | 6.01 | 5288.1 | 6570.1 | Positive | 19 |
| Patient 4 | 11.71 | 11.04 | 0.8 | 0.87 | 149.4 | 153.7 | Negative | 25 |
| Patient 5 | 6.47 | 6.58 | 5.29 | 5.44 | 5472.9 | 5468.1 | Positive | 20 |
| Patient 6 | 1.31 | 1.32 | 6.6 | 6.58 | > 40000.0 | > 40000.0 | Positive | 16 |
| Patient 7 | 14.16 | 13.51 | 6.8 | 6.98 | 12034.7 | 11191.5 | Positive | 22 |
| Patient 8 | 7.41 | 7.2 | 6.39 | 6.36 | 14489.9 | 12718.3 | Positive | 21 |
| Patient 9 | 15.15 | 14.86 | 4.73 | 4.82 | 2745.6 | 2603.7 | Positive | 20 |
| Patient 10 | 0.03 | 0.04 | 0.02 | 0.02 | 0.9 | 0.2 | Negative | 15 |
| Patient 11 | 1.27 | 1.22 | 0.22 | 0.23 | 186.4 | 186.8 | Negative | 20 |
| Patient 12 | 26.79 | 26.67 | 6.62 | 6.6 | 22392.8 | 19367.7 | Positive | 23 |
| Patient 13 | 5.81 | 6.06 | 4.31 | 4.33 | 1255.4 | 1183.3 | Positive | 19 |
| Patient 14 | 88.25 | 88.59 | 6.67 | 6.42 | 6811.4 | 6250 | Positive | 16 |
| Patient 15 | 88.27 | 88.14 | 6.66 | 6.58 | 6815.2 | 6587.5 | Positive | 21 |
| Patient 16 | 88.87 | 89.11 | 6.39 | 6.5 | 6924.4 | 6401.6 | Positive | 18 |
| Patient 17 | 2.09 | 2.2 | 6.23 | 6.19 | 1626.4 | 1630.5 | Positive | 17 |
| Patient 18 | 91.31 | 87.56 | 6.62 | 6.55 | 7459.3 | 6330 | Positive | 19 |
| Patient 19 | 4.56 | 4.58 | 7.36 | 7.31 | 2345.6 | 2274.9 | Positive | 19 |
| Patient 20 | 4.55 | 4.61 | 7.18 | 7.53 | 2491.1 | 2236.1 | Positive | 20 |

<sup>†</sup>An index value of $\geq 1.4$ is positive for IgG assays, $\geq 1.0$ is positive for IgM assays, a result of $\geq 50$ AU/ml per mL is positive for IgG II assays.

## 5. Discussion

Viral diversity will continue to challenge diagnostic tests as new SARS-CoV-2 strains arise and spread globally. Of particular interest are the B.1.1.7, B.1.351, and P.1 lineages that have been linked to increased transmissibility and immune escape [1-7]. *In silico* analyses did not predict impact to detection by Abbott molecular, antigen, and antibody assays; however, these predictions need to be continually tested. This study confirmed that Abbott COVID-19 molecular, antigen, and serological assays effectively detect SARS-CoV-2 B.1.1.7, B.1.351, and P.1 variant infections and B.1.1.7 antibodies.

In comparison to other RNA viruses, SARS coronaviruses maintain a higher rate of fidelity during replication due to an encoded proofreading mechanism [18]. Indeed, SARS-CoV-2 variants are typically defined by a pattern of mutations totaling <30 positions per genome (0.1% divergence), which is much lower than the divergence of 30% or more between HCV genotypes, for example [8, 19, 20]. In particular, antibody assays are less prone to impact due to the redundant nature of polyclonal immune responses as illustrated by the detection of spike antibodies in the sera of B.1.1.7 patients (Table 4). This observation is consistent with recent studies reporting that polyclonal antibodies in convalescent plasma bind the spike protein of SARS-CoV-2 B.1.1.7 and B.1.351 variants with similar binding kinetics despite variable capacity to neutralize the virus [21, 22]. Future studies should continue to explore the potential impact of SARS-CoV-2 sequence variation on diagnostic tests.

## Data Availability

All data referred to in the manuscript is shown in Tables presented in this pdf

## Acknowledgements

This study was funded by Abbott Laboratories.

